# Metabolic disorders and the risk of head and neck cancer: a protocol for a systematic review and meta-analysis

**DOI:** 10.1101/2021.10.14.21264986

**Authors:** Alexander Gormley, Charlotte Richards, Francesca Spiga, Emily Gray, Joanna Hooper, Barry G Main, Emma E Vincent, Rebecca C Richmond, Julian PT Higgins, Mark Gormley

**Author notes:** Corresponding author –, Mark Gormley, Bristol Dental School, University of Bristol, Lower Maudlin Street, Bristol, BS1 2LY. Joint first authors (Alexander Gormley, Charlotte Richards).

## Abstract

**Introduction:** Head and neck cancer squamous cell carcinoma (HNSCC) is the 6^th^ most common cancer internationally. Established risk factors include smoking, alcohol and presence of human papillomavirus (HPV). The incidence rate of new disease continues to rise, despite falls in alcohol consumption and a reduction in smoking, the rising rates are unlikely to be solely attributed to HPV status alone. Obesity and its associated conditions such as type 2 diabetes (T2D) are implicated in the risk and progression of a variety of cancers but there is paucity of evidence regarding its role in HNSCC.

**Methods and analysis:** A systematic review of cohort studies, reporting a risk of incident head and neck squamous cell carcinoma will be included. A systematic search strategy has been developed, multiple databases will be searched from January 1966, including Cochrane Library, OVID SP versions of Medline and EMBASE. The primary outcome will be incident HNSCC based on exposures of type 2 diabetes, obesity, dyslipidemia and hypertension as defined by the World Health Organisation (WHO). A combined risk effect across studies will be calculated using meta-analysis, although depending on the heterogeneity in study design, exposure and outcome reporting this may not be possible.

**Ethics and dissemination:** No ethical approval is required for this systematic review. The review will be published in a revelant peer-review journal and findings will be presented at scientific meetings in both poster and oral presentation form.

**Registration details:** This study has been registered with the international prospective register of systematic reviews (PROSPERO) with study registration number CRD42021250520. This protocol has been developed in accordance with the preferred reporting items for systematic review and meta-analysis protocols (PRISMA-P) guidance statement.

**Article Summary:** 

**Strengths and limitations of this study:**

- *This systematic review will be the first to comprehensively review the literature and provide a meta-analysis of the effect of metabolic disorders and the risk of incident HNSCC*.
- *The publication of this protocol provides a clear representation of the methods used in this review for transparency and to prevent future duplication*.
- *This systematic review will be one of the first to pilot the The ROBINS-E tool (Risk Of Bias In Non-randomised Studies - of Exposures)*.
- *Despite the metabolic disorders being described as separate entities, the authors recognise the potential for disease processes to be related*.
- *Among cohort studies there is significant variability in terms of length of follow-up for the outcome of interest, HNSCC*.

## Introduction

Head and neck squamous cell carcinoma (HNSCC), which includes cancer of the oral cavity, oropharynx and larynx, is the world’s 6th most common cancer, with the highest incidence in males and those over 70 years old.^1^ Established risk factors include smoking, alcohol and the human papillomavirus (HPV), which has mainly been linked to oropharyngeal cancer.^2^ Despite the significant reduction in smoking,^3^ and a fall in alcohol consumption,^4^ in the UK over the previous two decades, incidence rates of head and neck cancers have continued to rise by around a third.^1^ Given this potentially changing aetiology,^2^ further exploration of less well known risk factors is warranted to target prevention and identify patients with early disease. People living with obesity and associated conditions such as type 2 diabetes (T2D), have an increased risk of developing certain cancers (e.g. liver and pancreas), but the epidemiological evidence surrounding HNSCC is not conclusive.^5, 6^ Recent studies have demonstrated elevated glucose levels before or at the time of diagnosis, which may play a role in the pathogenesis and progression of cancer.^7^ Carcinogenesis in HNSCC is driven by diverse signalling pathways, however, little is known about the role of metabolism,^8^ despite metabolic reprogramming being a recognised hallmark of cancer.^9^

T2D is a common condition characterised by insulin resistance and hyperglycaemia, which results in whole body metabolic dysregulation.^10^ In 2015, the estimated worldwide diabetes prevalence for adults was predicted to rise to 642 million by 2040^11^. In addition, public health strategies have not successfully addressed the current ‘obesity pandemic’.^12-16^ One large pooled analysis demonstrated a weak positive association between T2D and HNSCC (OR 1.09; 95% confidence interval (95%CI): 0.95-1.24), which was stronger amongst those who never smoked cigarettes (OR 1.59; 95%CI: 1.22-2.07).^6^ While obesity is a potent risk factor for T2D, the relationship between body mass index (BMI) and HNSCC is not straightforward, with both positive and inverse associations observed.^17^ Another study found that a positive association with BMI was only observed in never smokers.^18^ In the largest study to date, waist circumference (WC) and waist-to-hip ratio (WHR) were positively associated with higher HNSCC risk, regardless of smoking status.^18^ Metabolic syndrome has been described as a clustering of disorders including obesity, hypertension, hyperglycaemia and dyslipidemia. A moderate inverse association has been observed between metabolic syndrome and HNSCC (OR 0.81; 95%CI, 0.78-0.85), but again these results were modified by tobacco use.^19^ Treatment for these conditions using medications such as statins,^20^ or metformin,^21^ may also reduce the risk of developing HNSCC. Given the growing pandemic of metabolic disorders, understanding how these alterations affect the risk of carcinogenesis may identify those who are high risk. This could drive targeted prevention, earlier detection and perhaps the identification or repurposing of drug targets for treatment in HNSCC.^22^

### Rationale

The complex metabolic changes associated with T2D, obesity, dyslipidemia and hypertension may alter the risk of certain cancers, but the evidence for head and neck cancer is inconclusive. This study aims to identify, collate and synthesise all relevant studies including adult participants, to determine whether the risk of developing incident HNSCC is influenced by T2D, obesity, dyslipidemia and hypertension, using relevant effect measures (e.g., odds or risk ratios). Where available, we will further stratify by subsite (i.e., oral cavity, oropharynx and larynx), as well as by HPV status.

### Methods

#### Research Question

‘Do metabolic disorders affect the risk of developing head and neck cancer?’

#### Study Design

Randomised controlled trials are informative for assessing causal effects, however due to the nature of the exposures in this study, inclusion of this design are not feasible. Therefore, we will focus on observational (e.g., cohort studies) reporting the risk of metabolic disorders on incident head and neck squamous cell carcinoma.

#### Eligibility criteria

**Population**-participants over 18 years old, of either sex, from any ethnic background.

**Exposures-** 1) type 2 diabetes, 2) obesity, 3) dyslipidemia and 4) hypertension. These will be collectively described as ‘metabolic disorders.

**Comparison**-participants who have not been diagnosed with the aforementioned metabolic disorders.

**Outcome**-incident head and neck squamous cell carcinoma.

The study will report in line with the Preferred Reporting Items for Systematic Reviews and Meta-Analyses (PRISMA) statement and has been pre-registered on International Prospective Register of Systematic Reviews (PROSPERO) in 2021 (CRD42021250520).

#### Search strategy

A systematic search strategy (**Supplementary Table 1**) has been formulated by clinicians, scientific researchers and a specialist librarian for Surgery, Head & Neck and Medicine. Medical Subject Headings (MeSH) and keywords will be iteratively combined with the Boolean operators AND, OR and NOT. Multiple databases will be searched from January 1966, including Cochrane Library, OVID SP versions of Medline and EMBASE. Pre-print servers including medRxiv and bioRxiv will also be searched. In addition, the following electronic bibliographic databases will be searched: EThOS, Google Scholar, Open Grey and ClinicalTrials.gov, to identify articles from the grey literature and conference proceedings. extracted from the full-length articles will be reviewed to identify other publications of interest. Duplicate articles will be removed using Covidence©^23^.

#### Data management

The results from searches will be imported into Covidence© software, to improve reference management and workflow. This will also populate a PRISMA flow diagram.^24^

#### Data screening and extraction

All titles and abstracts will be screened by two authors A.G. and C.R., with conflicts resolved by A.G., C.R. and M.G. Screening records and decisions will be kept in Covidence©. Data will be extracted from titles that meet the inclusion criteria (**Table 1**). The data extraction form (**Supplementary Table 2**) will be piloted using 5 included titles with A.G., C.R. and M.G. independently completing data extraction. If amendments are necessary they will be performed at this point, with consensus approval required prior to final use of the form. Subsequent to this, C.R. and M.G. will double data extract indepdently for each title, any disagreements will be discussed between A.G., C.R. and M.G. with consensus approval required.

**Table 1.**
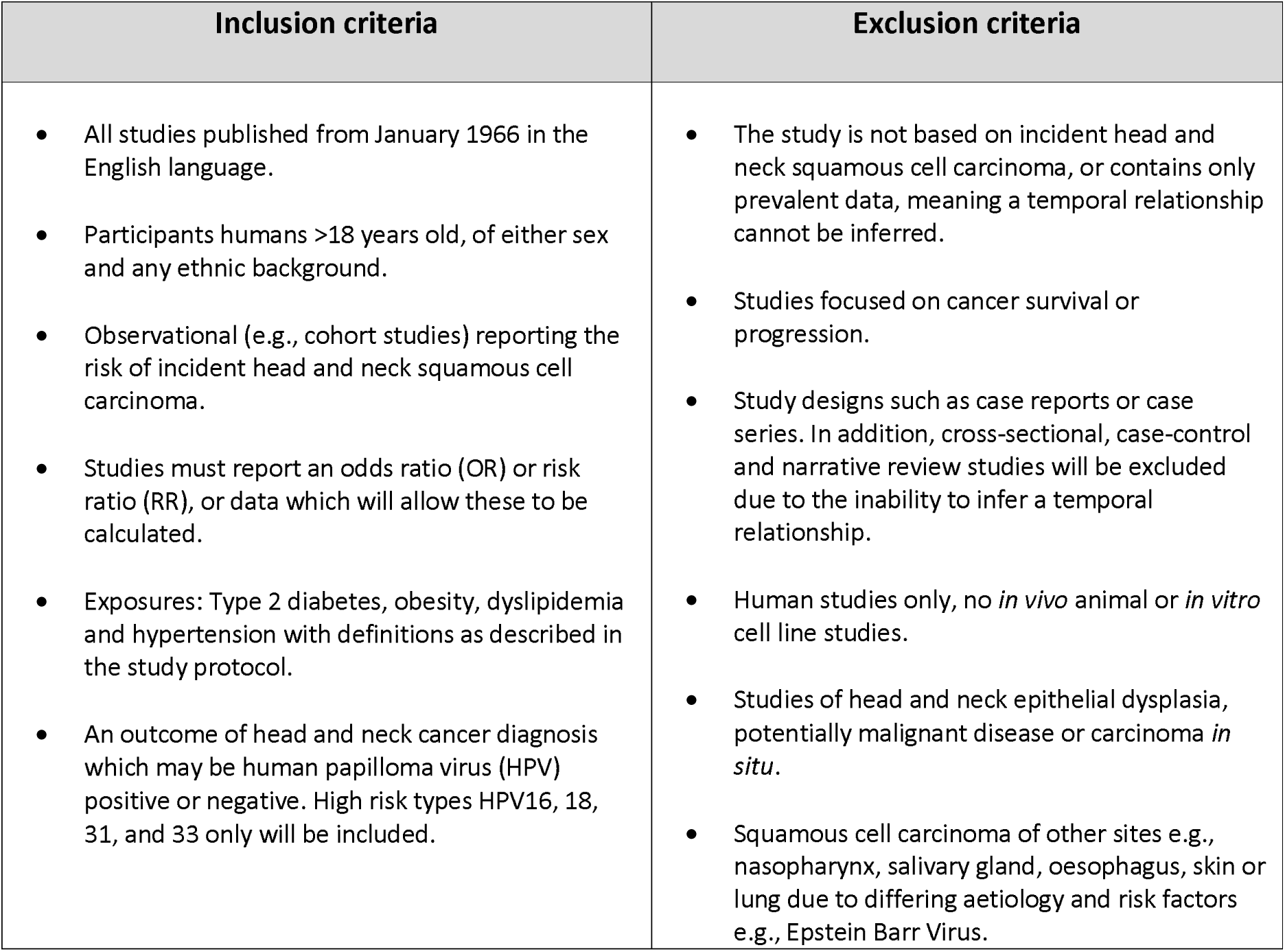
Study selection criteria

| Inclusion criteria | Exclusion criteria |
| --- | --- |
| All studies published from January 1966 in the English language.<br>Participants humans >18 years old, of either sex and any ethnic background.<br>Observational (e.g., cohort studies) reporting the risk of incident head and neck squamous cell carcinoma.<br>Studies must report an odds ratio (OR) or risk ratio (RR), or data which will allow these to be calculated.<br>Exposures: Type 2 diabetes, obesity, dyslipidemia and hypertension with definitions as described in the study protocol.<br>An outcome of head and neck cancer diagnosis which may be human papilloma virus (HPV) positive or negative. High risk types HPV16, 18, 31, and 33 only will be included. | The study is not based on incident head and neck squamous cell carcinoma, or contains only prevalent data, meaning a temporal relationship cannot be inferred.<br>Studies focused on cancer survival or progression.<br>Study designs such as case reports or case series. In addition, cross-sectional, case-control and narrative review studies will be excluded due to the inability to infer a temporal relationship.<br>Human studies only, no *in vivo* animal or *in vitro* cell line studies.<br>Studies of head and neck epithelial dysplasia, potentially malignant disease or carcinoma *in situ*.<br>Squamous cell carcinoma of other sites e.g., nasopharynx, salivary gland, oesophagus, skin or lung due to differing aetiology and risk factors e.g., Epstein Barr Virus. |

### Disease definitions

1. The World Health Organization (WHO) defines T2D as a chronic disease that occurs when the body cannot effectively use the insulin it produces and is largely the result of excess body weight and physical inactivity. A diagnosis of T2D is defined as symptoms such as polyuria or polydipsia, plus:^25, 26^
  - a random blood plasma glucose concentration ≥ 11.1 mmol/l or;
  - a fasting plasma glucose concentration ≥ 7.0 mmol/l (whole blood ≥ 6.1 mmol/l) or;
  - two hour plasma glucose concentration ≥ 11.1 mmol/l two hours after 75g anhydrous glucose in an oral glucose tolerance test (OGTT) or;
  - glycated haemoglobin (HbA1c) 6.5% or more (48 mmol/mol and above)
2. The World Health Organization (WHO) defines obesity as having: a body mass index (BMI) of 30 or above.^27^
3. Dyslipidemia is classified as serum total cholesterol (TC), low density lipoprotein cholesterol (LDL-C), triglycerides, apolipoprotein B, or lipoprotein(a) concentrations above the 90^th^ percentile, or high density lipoprotein cholesterol (HDL-C) or apolipoprotein concentrations below the 10th percentile for the general population.^28^
4. The World Health Organization (WHO) defines hypertension as:^29^
  - a systolic blood pressure reading of ≥140 mmHg when recorded on two different days; and/or
  - a diastolic blood pressure readings on both days of ≥90 mmHg.

A diagnosis of incident head and neck squamous cell carcinoma should be confirmed by histology by a trained pathologist. Cancer diagnosis and confirmation may be reported using the International Classification of Diseases (ICD) codes.^30^ Cancer cases for the disease of interest have the following ICD codes: oral cavity (C02.0-C02.9, C03.0-C03.9, C04.0-C04.9, C05.0-C06.9) oropharynx (C01.9, C02.4, C09.0-C10.9), hypopharynx (C13.0-C13.9), overlapping (C14 and combination of other sites) and 25 cases with unknown ICD code (other). With respect to HPV diagnosis, only high risk subtypes HPV16, 18, 31, and 33 will be included. These are should be detected using HPV-DNA via polymerase chain reaction (PCR) and/or in situ hybridization (ISH) examination, usually used in combination with immunohistochemistry for *p16*.

### Data analysis and synthesis

The analysis and synthesis process will be performed in two phases. The first phase consists of estimating the effect of each metabolic disorder (T2D, obesity, dyslipidemia and hypertension) on incident HNSCC separately for each included study. Effects should be reported as odds (OR) or risk ratios (RR), with 95% confidence intervals or in a format that will enable us to calculate these.

The second phase will focus on estimating the combined effect across studies using meta-analysis, using metafor software in R (R Core Team, version 4.0.3).^31^ The analysis will yield information about the heterogeneity of effects across studies, using Cochran’s Q test and Higgins’ I^2^ statistic. Due to the heterogeneity in study design, exposure definition and outcome reporting in the included studies, meta-analysis may not be reasonable.

### Subgroup analyses

Where possible, we will stratify by oral, oropharyngeal and laryngeal subsite and by HPV-status as post-hoc analyses, to determine if effects seen are specific to a particular anatomical region or associated with specific tumour types.

### Risk of bias in studies

Two authors will independently extract the relevant risk factor information during data extraction using a preliminary version of the Risk of Bias in Non-randomised Studies – of Exposures (ROBINS-E) tool (https://www.bristol.ac.uk/population-healthsciences/centres/cresyda/barr/riskofbias/robins-e/).^32^ The ROBINS-E tool aims to assess the result of interest from an observational study for risk of bias, where that study is designed to assess the effect of exposure on outcome.^32^ Disagreements will be recorded and discussed with a third author for resolution. This assessment will be used in evaluating the strength of evidence from the included studies.

### Amendments

Any amendments to the initial protocol will be included in an update to the PROSPERO record, including information on amendment type, reasoning and a timestamp. Deviation from this protocol will be described in the full systematic review paper.

## Supporting information

PRISMA-P Checklist

## Data Availability

This is a protocol for a systematic review where no primary data will be collected, the search strategy is included in the supplementary data for where the data for this systematic review will be obtained from.

## Dissemination

This systematic review will be published in a relevant peer-reviewed journal and findings from this work may be presented at scientific meetings (both poster and oral presentations).

## Ethical approval

No ethical approval is required for the conducting of this systematic review.

## Acknowledgements

None to declare.

## Authors’ contributions

AG and CR jointly contributed to the development of the protocol and the drafting, writing and editing of this manuscript. FS contributed to the development of this study and editing of the manuscript. JH contributed to the development of the search strategy. BM, EG, EV, RR and JPTH contributed to the development of this study. MG was responsible for conceptualising this study, drafting and editing this manuscript. All authors contributed to the development of the search strategy. All authors have approved and contributed to the final written manuscript.

## Competing interests statement

None declared

**Supplementary Table 1.**
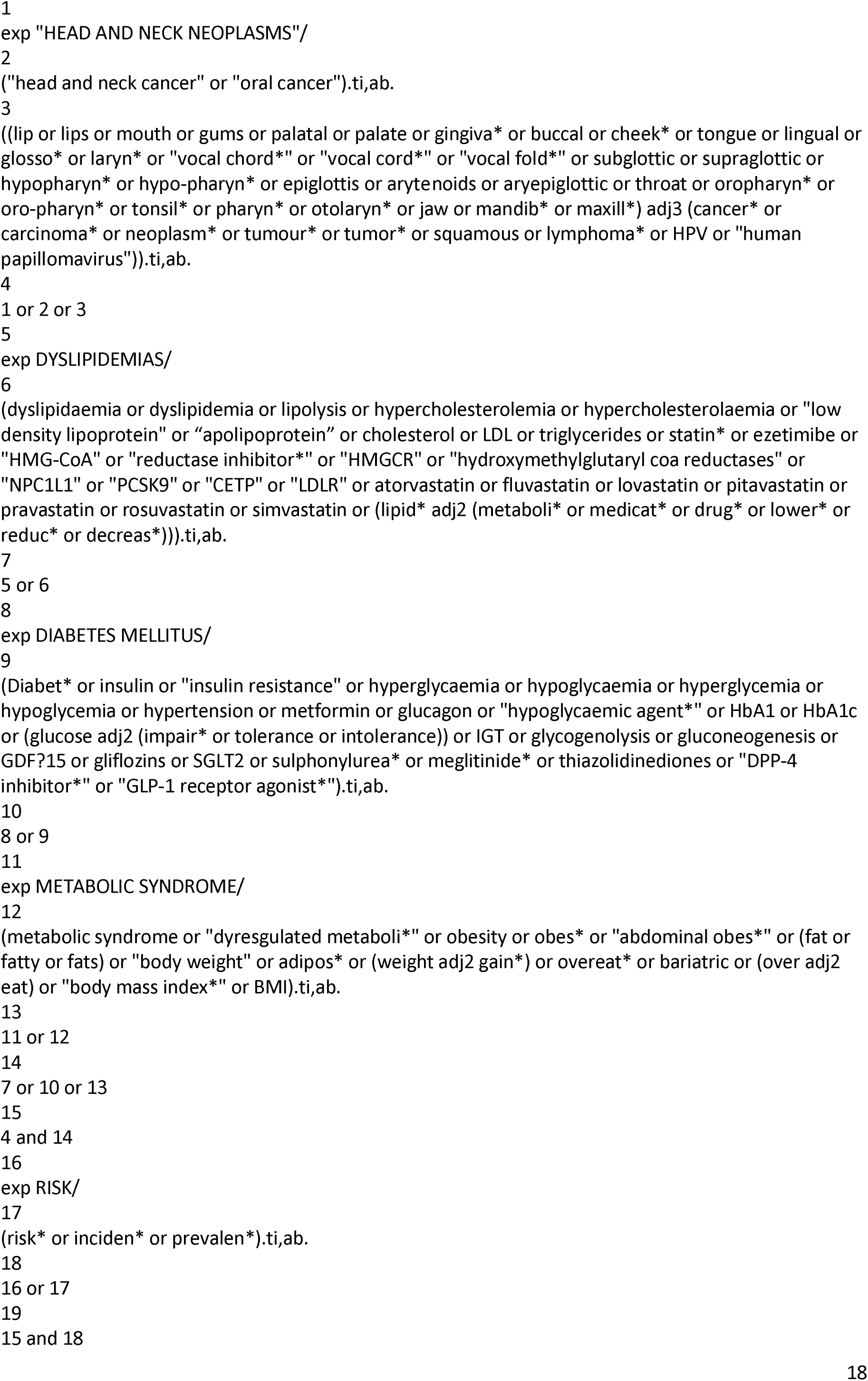
Search strategy

**Supplementary Table 2.**
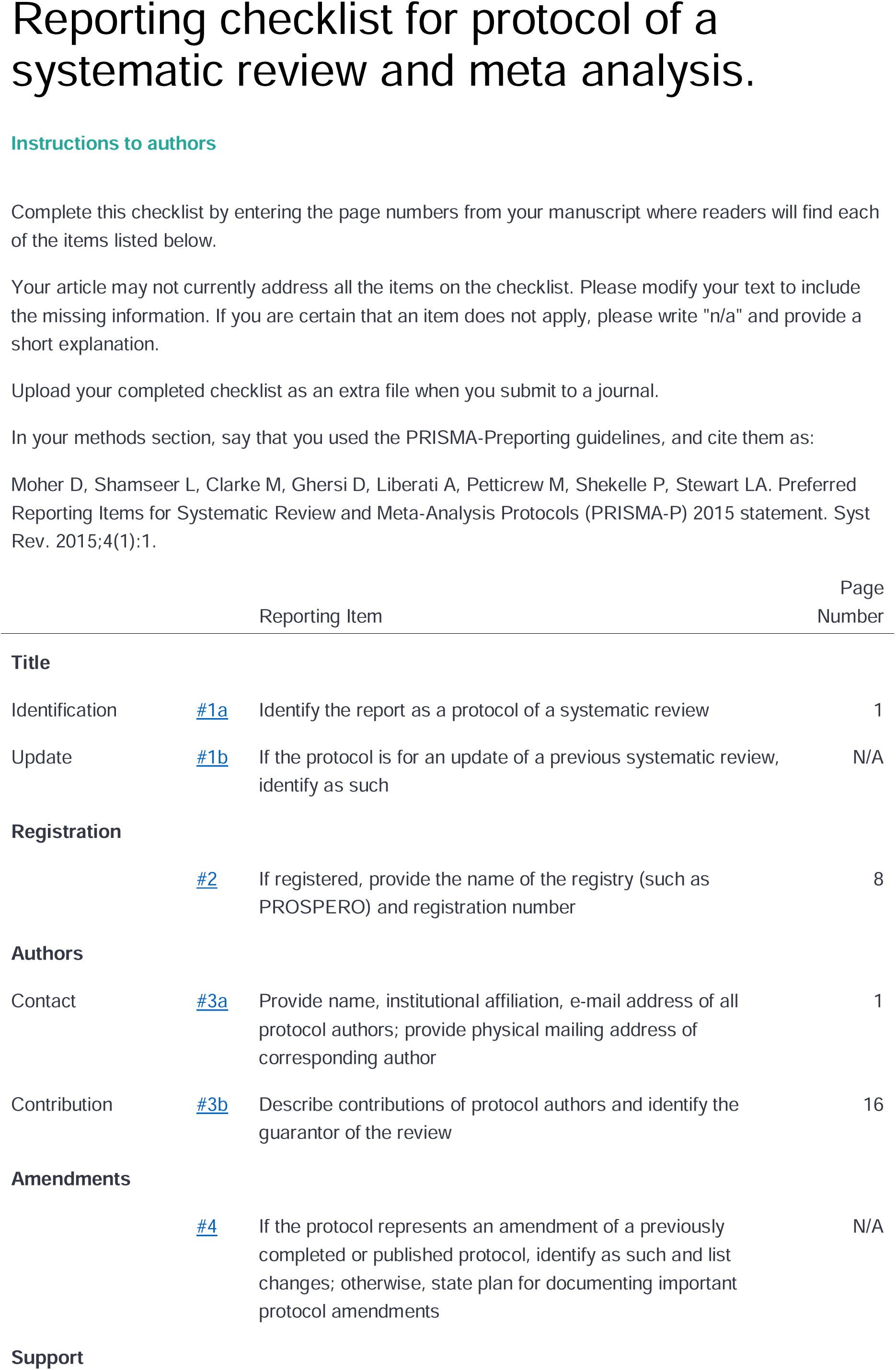
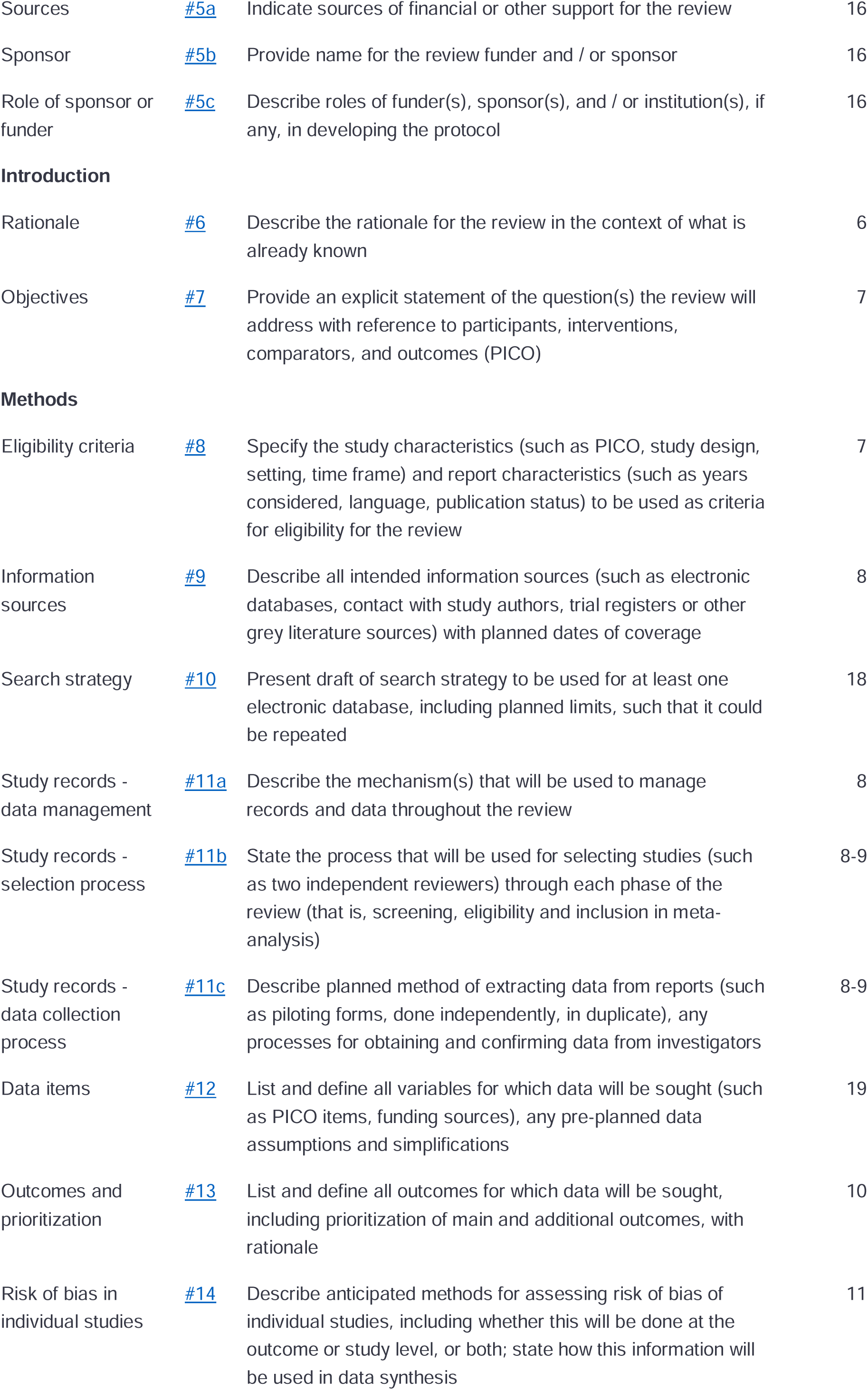
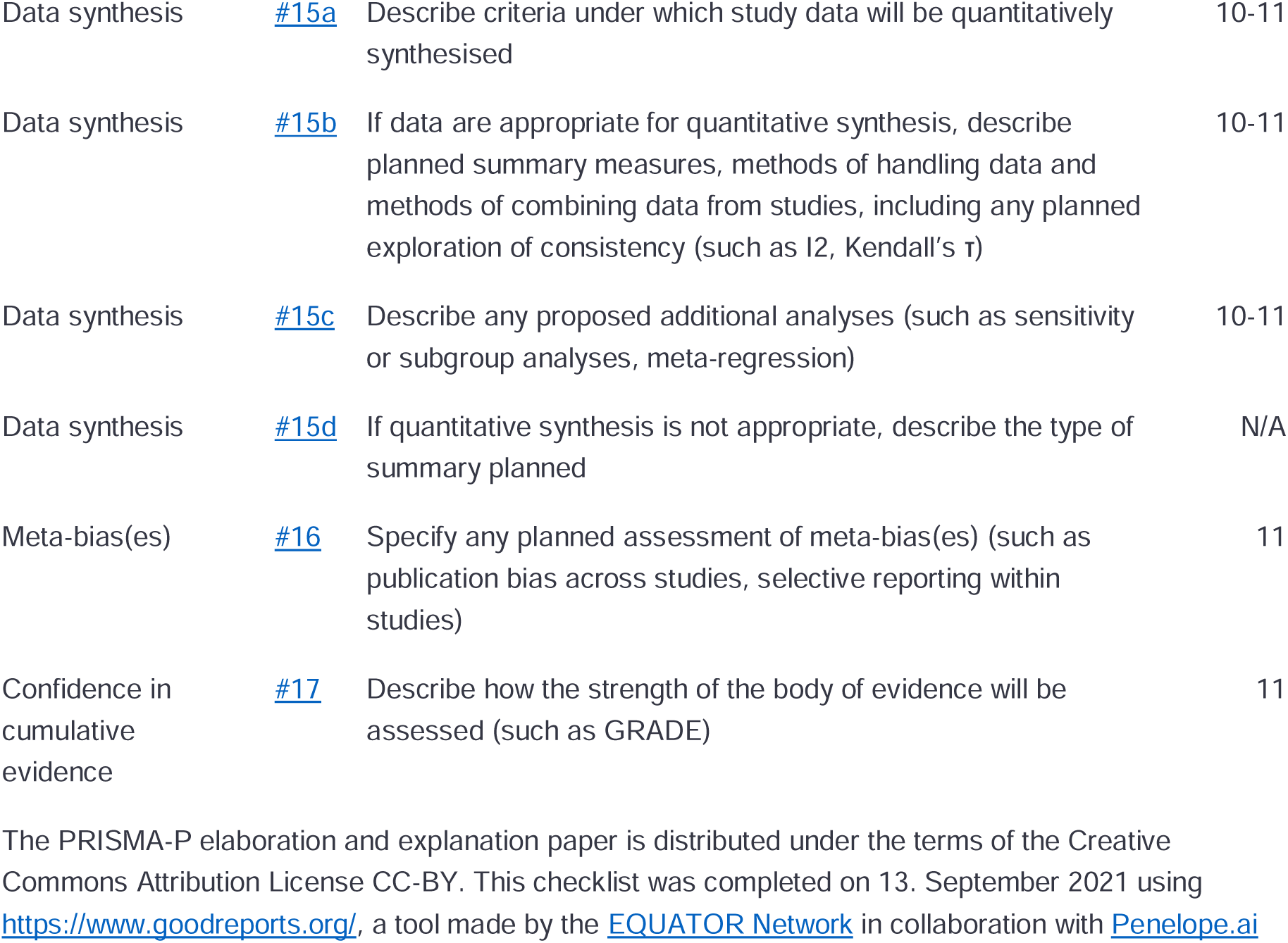
Data extraction table. Note separate table for each exposure (Obesity, Dyslipidaemia, Hypertension, Type 2 diabetes).

